# Single-cell RNAseq identifies clonally expanded antigen-specific T-cells following intradermal injection of gold nanoparticles loaded with diabetes autoantigen in humans

**DOI:** 10.1101/2023.07.05.23291245

**Authors:** Stephanie J. Hanna, Terri C. Thayer, Emma J.S. Robinson, Ngoc-Nga Vinh, Nigel Williams, Laurie Landry, Robert Andrews, Qi Zhuang Siah, Pia Leete, Rebecca Wyatt, Martina A. McAteer, Maki Nakayama, F. Susan Wong, Jennie H.M. Yang, Tim I.M. Tree, Johnny Ludvigsson, Colin M. Dayan, Danijela Tatovic

**Author notes:** Corresponding author: Dr Danijela Tatovic, Division of Infection and Immunity, Cardiff University School of Medicine, Tenovus Building, Heath Park, Cardiff, CF14 4XN. Shared senior authorship (Colin M. Dayan, Danijela Tatovic).

## Abstract

Gold nanoparticles (GNPs) have been used in the development of novel therapies as a way of delivery of both stimulatory and tolerogenic peptide cargoes. Here we report that intradermal injection of GNPs loaded with the proinsulin peptide C19-A3, in patients with type 1 diabetes, results in recruitment and retention of immune cells in the skin. These include large numbers of clonally expanded T-cells sharing the same paired T-cell receptors (TCRs) with activated phenotypes, half of which, when the TCRs were re-expressed in a cell-based system, were confirmed to be specific for either GNP or proinsulin. All the identified gold-specific clones were CD8^+^, whilst proinsulin-specific clones were both CD8^+^ and CD4^+^. Proinsulin-specific CD8^+^ clones had a distinctive cytotoxic phenotype with overexpression of granulysin (GNLY) and KIR receptors. Clonally expanded antigen-specific T cells remained *in situ* for months to years, with a spectrum of tissue resident memory and effector memory phenotypes.

As the T-cell response intradermally is divided between targeting the gold core and the antigenic cargo, this offers a route to improving Trm formation in response to vaccines. In addition, our scRNAseq data indicate that focussing on clonally expanded skin infiltrating T-cells recruited to intradermally injected antigen is a highly efficient method to enrich and identify antigen-specific cells. This approach has the potential to be used to monitor the intradermal delivery of antigens and nanoparticles for immune modulation in humans.

## INTRODUCTION

Antigen-specific immunotherapy (ASI) for Type 1 diabetes (T1D) aims to abrogate the immune response to β-cell antigens, without systemic dampening of the immune system. In a Phase 1 safety study to assess the use of ultra-small (< 5nm) gold nanoparticles (GNPs) for this indication, we conjugated the proinsulin peptide C19-A3 to the gold core via a thiol linker and injected these C19-A3 GNPs intradermally using a clinically approved microneedle delivery system, MicronJet600. We have reported elsewhere on the preclinical data associated with this formulation(1) and the clinical results of the trial(2). Proinsulin peptide C19-A3 has been reported to be HLA-DR4 (DRB1*0401) restricted and, given intradermally, with or without GNP, has been shown to be safe(2, 3). When given alone, it demonstrated the potential for tolerance induction in recipients with T1D(4). *In vitro* experiments demonstrated that conjugation of diabetogenic peptides to GNPs facilitated their uptake by antigen presenting cells, particularly Langerhans cells in the skin(1), which have been shown to have tolerogenic properties in some studies(5, 6). In the Phase 1 trial, five participants with newly diagnosed T1D received multiple rounds of C19-A3-GNP and we found persistent reactive immune infiltrate at the C19-A3 GNP injection site(2).

This provided the unique opportunity to analyse the antigen specificity and phenotype of the tissue infiltrate, rather than the historical approach of peripheral blood analysis, where autoantigen-specific cells are very rare, representing ∼0.01% of the total T-cell population(7–9). Consequently, the characterisation of these cells is challenging and represents a barrier to monitoring antigen-specific T-cell phenotypes during diabetes development, progression, and treatment with novel immunotherapies(10). Much work on antigen-specific cells in T1D has used tetramers to isolate antigen-specific cells from the peripheral blood(8), but is limited by HLA restriction and to known antigen epitopes(11). An alternative approach has been to use *in vitro* stimulation of cells from the peripheral blood with whole diabetes autoantigens or peptides(7, 12, 13) and these studies have suggested that diabetes autoantigen epitopes and HLA restriction exhibit considerable diversity. However, issues of low cell frequency remain with these assays.

In other autoimmune diseases, many insights have come from extraction of lymphocytes from the site of disease, such as from the synovium in psoriatic arthritis. These cells of predominantly memory phenotype are also found in the blood of affected individuals(14). In T1D, whilst antigen-specific lymphocytes are enriched in the pancreas and pancreatic lymph node, these tissues are inaccessible, and thus only suitable for analysis in deceased donors. However, it has been demonstrated that T-cells, particularly CD8^+^ T-cells, that infiltrate the pancreatic islets in T1D are highly clonal(15–19).

Here we report the characterisation of tissue immune cell responses following intradermal injection of C19-A3 GNP, using a state-of-the art method of single cell RNA sequencing (scRNAseq)(16).

## RESULTS

### Study participants

A total of five participants who received intradermal microneedle injection of C19-A3 GNP were included in the final analysis. Whilst four participants received 3 doses of C19-A3 GNP, one participant (EEASI-C) received only the first dose due to the decision to halt further drug administration, but they remained in the study and attended all planned visits and assessments. All participants were autoantibody-positive defined by being positive to at least one of GADA, IA-2A, or ZnT8A and possessed HLA-DRB1∗0401 genotype(2). Two participants had a punch biopsy of the injection sites, 3 months after injection in one (participant EEASI-B) and 7 months after injection in another participant (participant EEASI-A). Three participants had a suction blister raised over the injection sites: 10 months after injection in two participants (participants EEASI-B and EEASI-C) and 28 months after injection in the third participant (participant EEASI-A).

### Intradermal injection of C19-A3 GNP results in sustained GNP deposits in the skin and immune cell infiltration

As previously reported(2), C19-A3 GNP injections induced formation of a dark-coloured bleb (diameter 3–4 mm) (Figure 1a) in all participants, with transient surrounding erythema lasting up to 60 min post injection. The dark area persisted and reduced in size to 1–2 mm by the following day. Subsequently, delayed skin changes over the injection site were observed. The change consisted of a small central area of GNP-derived pigmentation surrounded by persistent asymptomatic circular erythematous induration. Induration initially reached 2–3 cm in diameter, which then reduced to approximately 1 cm during the follow-up period. These delayed reactions appeared 17.4 ± 2.3 days after the first injection, but much sooner, i.e. 1– 3 days after the second and third injection, a reaction that was consistent in all participants. The skin changes faded significantly over the course of the study, although they were still visible at the end of the observation period (12–24 months depending on the participant).

**Figure 1.**
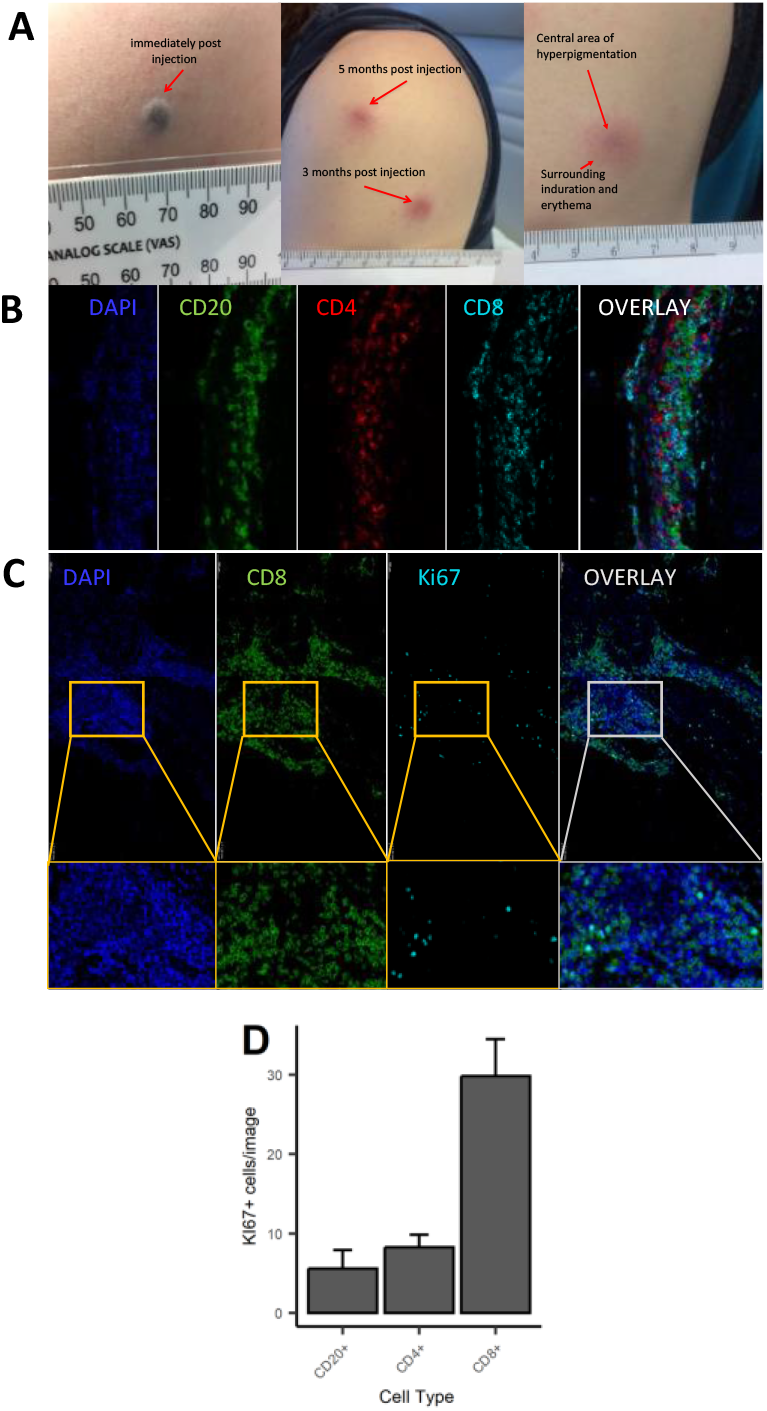
Macroscopic and microscopic appearance of skin changes following intradermal injection of C19-A3 GNP. A) Images show representative injection sites in deltoid region immediately after injection (left), several months after injection (centre) with the enlarged image showing dark centre and surrounding erythematous induration (right); B,C) Sections from punch skin biopsies were stained for CD20, CD4, CD8 and Ki-67 and imaged with a fluorescence confocal microscope. CD8^+^ T Lymphocytes predominated in all parts of the immune clusters: total overall counts/1.54mm^2^ area: CD8^+^=6537, CD4+=1526, CD20^+^=1363/1.54mm^2^; D) number of Ki67^+^ cells was higher amongst CD8^+^ cells vs CD20^+^ and CD4^+^ cells.

A skin biopsy of the injection site was performed on two participants (3 months after injection in one (participant EEASI-B) and 7 months after injection in another participant (participant EEASI-A)). Confocal microscopy showed that CD8^+^ T-cells predominated in all parts of the immune clusters (Figure 1B and C) with total overall counts as follows CD8^+^=6537, CD4^+^=1526, CD20^+^=1363/1.54mm^2^ area. CD8^+^ T-cells proportionally contained higher numbers of Ki67^+^ cells in comparison to CD20^+^ and CD4^+^ cells (Figure 1D). There was also evidence of intradermal gold retention as previously reported(2).

To examine the characteristics of the cells that were involved in the injection site reaction, we formed suction blisters at the injection site in three participants and performed scRNAseq and TCR VDJ sequencing on the cells in the blister fluid. After quality control and doublet removal, cells were visualised using a UMAP projection (Figure 2A). Cell types identified included memory CD8^+^ T-cells, memory CD4^+^ T-cells, Tregs, dendritic cells, melanocytes and keratinocytes. The findings were consistent across all three participants (Figure 2B).

**Figure 2.**
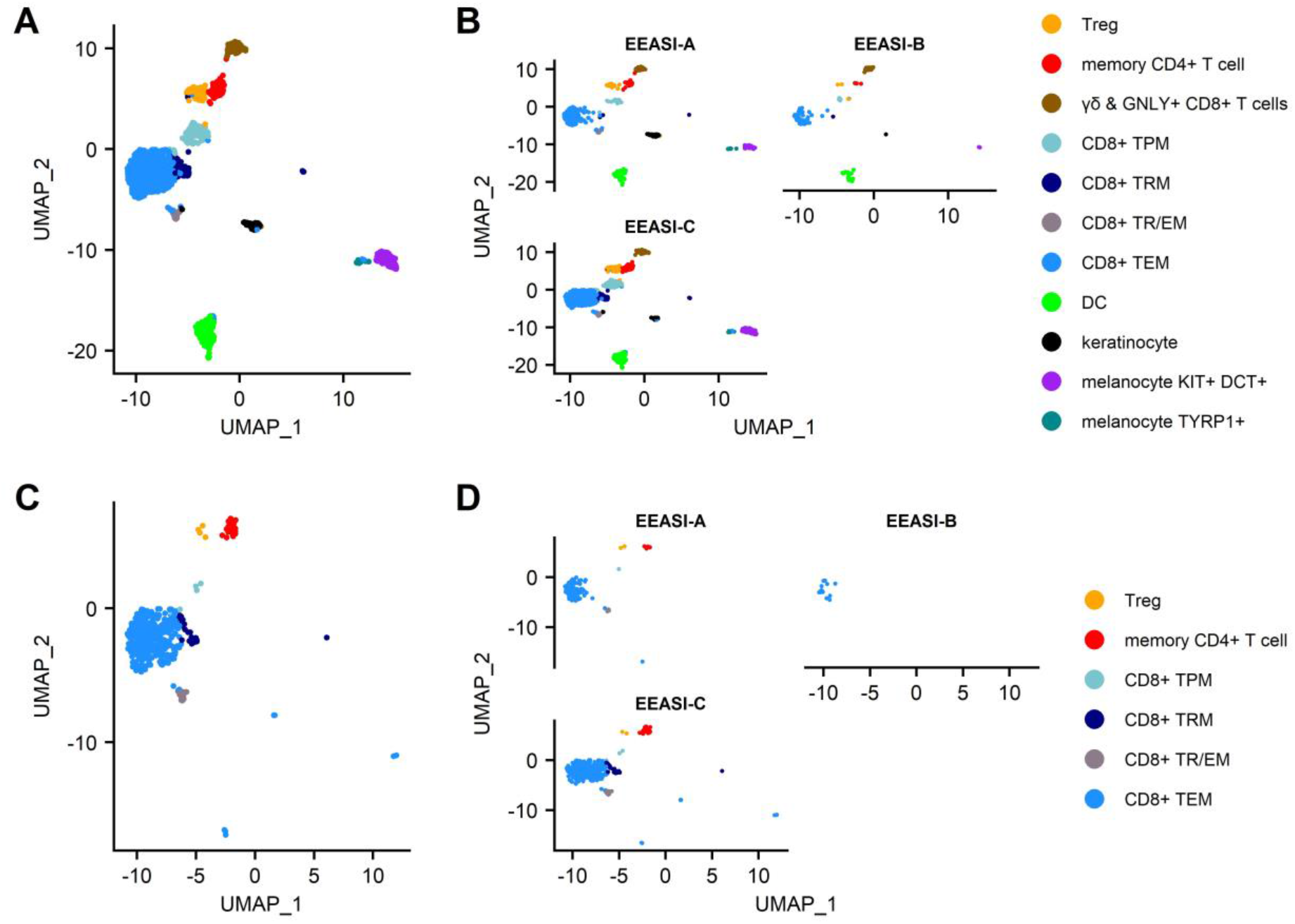
scRNAseq reveals memory T-cell infiltration of injection site of C19-A3 GNP and clonal expansions. A) UMAP plot of pooled samples of cells from the blister fluid of 3 donors (B) UMAP plots of the blister fluid cells split by donor; C) UMAP plot of all clonally expanded T-cells in the blisters (clones share an identical TCRα and TCRβ CDR3, or where one chain is not detected, the remaining chain is identical); D) UMAP plot of clonally expanded T-cells in the blister split by donor.

The cell types labelled in the UMAP projections were those that were clustered automatically using Seurat, with the exception of the CD4^+^ memory T-cells and Treg cells, which were initially a single cluster. This cluster was manually gated into two groups and the top 10 DEGs were used to discern the separate phenotypes. This revealed a distinct FOXP3^+^, CTLA4^+^, TIGIT^+^ population of Tregs, whereas the other CD4^+^ cells had a memory phenotype (Supplementary Figure 1A). The remaining CD8^+^ T-cells clustered into five groups and were defined using known phenotypic markers: 1. T_EM_ – T-cell effector memory comprised the largest subset that expressed CD69, granzyme B (GzB), perforin. They also expressed PD-1; 2. T_RM/EM_ – a subset of true tissue resident T-cells with some characteristics of effector memory, which express CD103 and CD69. They are proliferating (KI67^+^) and are CXCR3^+^, and GZB^+^; 3. T_RM_-T-cell resident memory that expressed CD69, CD27 and PD1, but did not demonstrate cytotoxic gene expression; 4. T_PM_ - peripheral memory T-cells that express CD69 but also CD62L, CCR7 and CD127. They lack cytotoxic or cytokine gene expression; 5. γδ and Granulysin (GNLY)^+^ CD8^-^ highly cytotoxic effector memory CD69^+^, expressing GNLY, GzB, perforin. They are CD127^+^, and express XCL1 and CXCR3. (Supplementary Figure 1). B cells were not detected in the blister fluid, despite being abundant in the punch biopsies and this was likely to be due to a reduced ability to migrate into the fluid in the timescale of the blister formation.

### Single cell RNAseq analysis of T-cells harvested from the injection site reveals significant clonal expansions specific for the injected antigens

We examined the sequences of the paired α-and β-chains of the TCRs for each T-cell (TCR clonotype). In addition, there were cells that shared the TCRβ with other clones but lacked a sequenced α-chain as the mRNA for the α-chain is less abundant in T-cells(20). A striking feature was the presence of a large number of clonally expanded T-cells sharing the same paired TCRs. These clonal expansions were frequently very large. In the case of EEASI-C the largest expansion comprised 52 clonally identical cells, out of a total of 807 cells with sequenced TCRs (Figure 2D and Table 1). The phenotypes of the clonally expanded T-cells included memory CD4^+^ T-cells, Tregs, CD8^+^T_TR/EM_, CD8^+^T_EM_, CD8^+^T_PM_, and CD8^+^T_RM_ (Figure 2C and D) but not γδ and GNLY+ CD8^+^ T-cells. Two of the three participants had clones falling into all compartments listed above. However, EEASI-B had only CD8^+^T_EM_ expanded clones (Figure 2D).

**Table 1.**
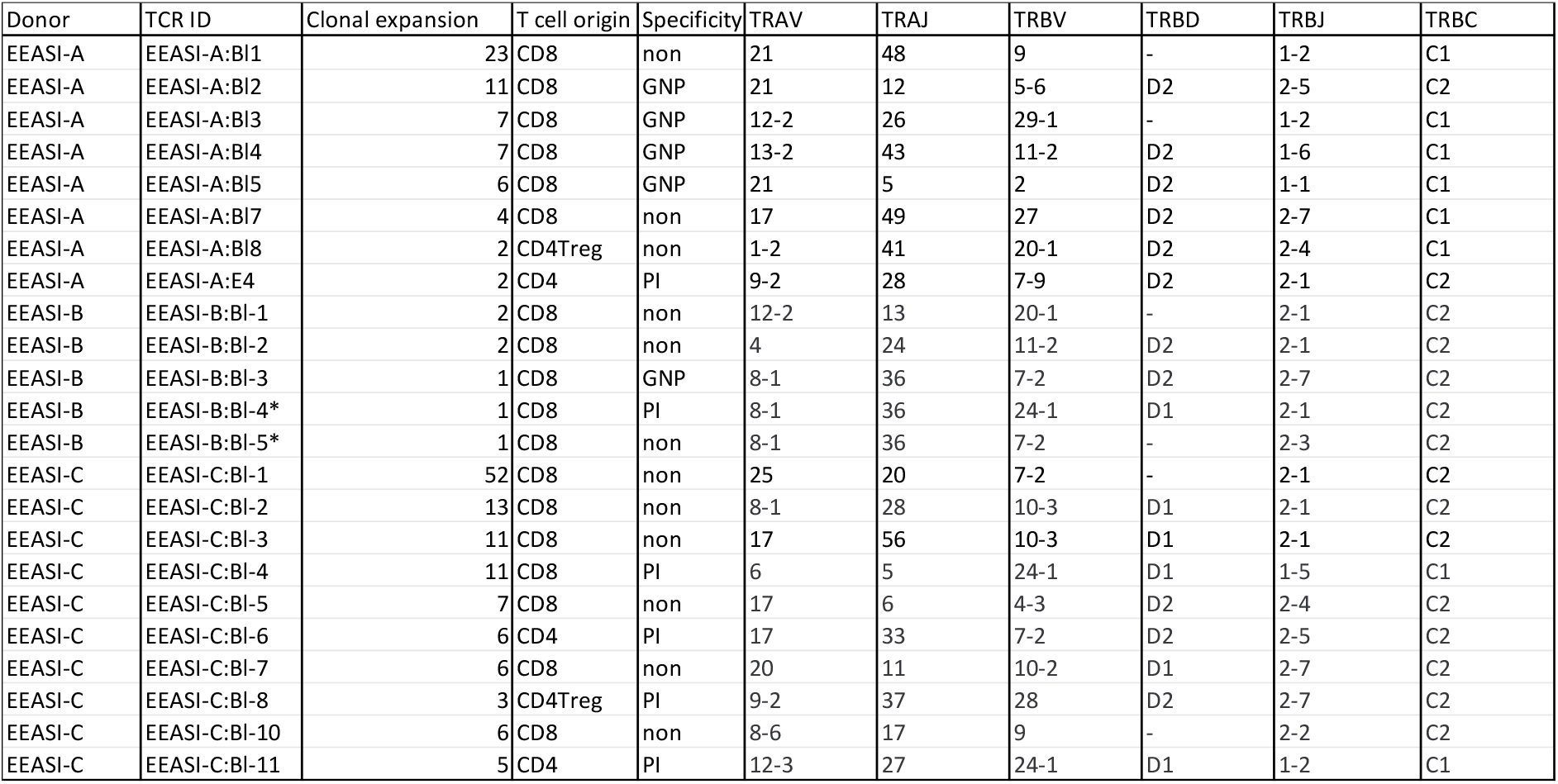
Phenotype, reactivity, TCR sequences and number of copies of the clones harvested from the blister fluid raised over the C19-A3 GNP injection sites. * In general, sequenced T-cells had a single TCRα and TCRβ, however, EEASI-B:Bl-4 and EEASI-B:Bl-5 come from a cell with two TCRβ-chains and a single TCRα-chain remade as two separate 5KC cell lines. Non denotes non-specific for either PI or GNP.

| Donor | TCR ID | Clonal expansion | T cell origin | Specificity | TRAV | TRAJ | TRBV | TRBD | TRBJ | TRBC |
| --- | --- | --- | --- | --- | --- | --- | --- | --- | --- | --- |
| EEASI-A | EEASI-A:BI1 | 23 | CD8 | non | 21 | 48 | 9 | - | 1-2 | C1 |
| EEASI-A | EEASI-A:BI2 | 11 | CD8 | GNP | 21 | 12 | 5-6 | D2 | 2-5 | C2 |
| EEASI-A | EEASI-A:BI3 | 7 | CD8 | GNP | 12-2 | 26 | 29-1 | - | 1-2 | C1 |
| EEASI-A | EEASI-A:BI4 | 7 | CD8 | GNP | 13-2 | 43 | 11-2 | D2 | 1-6 | C1 |
| EEASI-A | EEASI-A:BI5 | 6 | CD8 | GNP | 21 | 5 | 2 | D2 | 1-1 | C1 |
| EEASI-A | EEASI-A:BI7 | 4 | CD8 | non | 17 | 49 | 27 | D2 | 2-7 | C1 |
| EEASI-A | EEASI-A:BI8 | 2 | CD4Treg | non | 1-2 | 41 | 20-1 | D2 | 2-4 | C1 |
| EEASI-A | EEASI-A:E4 | 2 | CD4 | PI | 9-2 | 28 | 7-9 | D2 | 2-1 | C2 |
| EEASI-B | EEASI-B:BI-1 | 2 | CD8 | non | 12-2 | 13 | 20-1 | - | 2-1 | C2 |
| EEASI-B | EEASI-B:BI-2 | 2 | CD8 | non | 4 | 24 | 11-2 | D2 | 2-1 | C2 |
| EEASI-B | EEASI-B:BI-3 | 1 | CD8 | GNP | 8-1 | 36 | 7-2 | D2 | 2-7 | C2 |
| EEASI-B | EEASI-B:BI-4* | 1 | CD8 | PI | 8-1 | 36 | 24-1 | D1 | 2-1 | C2 |
| EEASI-B | EEASI-B:BI-5* | 1 | CD8 | non | 8-1 | 36 | 7-2 | - | 2-3 | C2 |
| EEASI-C | EEASI-C:BI-1 | 52 | CD8 | non | 25 | 20 | 7-2 | - | 2-1 | C2 |
| EEASI-C | EEASI-C:BI-2 | 13 | CD8 | non | 8-1 | 28 | 10-3 | D1 | 2-1 | C2 |
| EEASI-C | EEASI-C:BI-3 | 11 | CD8 | non | 17 | 56 | 10-3 | D1 | 2-1 | C2 |
| EEASI-C | EEASI-C:BI-4 | 11 | CD8 | PI | 6 | 5 | 24-1 | D1 | 1-5 | C1 |
| EEASI-C | EEASI-C:BI-5 | 7 | CD8 | non | 17 | 6 | 4-3 | D2 | 2-4 | C2 |
| EEASI-C | EEASI-C:BI-6 | 6 | CD4 | PI | 17 | 33 | 7-2 | D2 | 2-5 | C2 |
| EEASI-C | EEASI-C:BI-7 | 6 | CD8 | non | 20 | 11 | 10-2 | D1 | 2-7 | C2 |
| EEASI-C | EEASI-C:BI-8 | 3 | CD4Treg | PI | 9-2 | 37 | 28 | D2 | 2-7 | C2 |
| EEASI-C | EEASI-C:BI-10 | 6 | CD8 | non | 8-6 | 17 | 9 | - | 2-2 | C2 |
| EEASI-C | EEASI-C:BI-11 | 5 | CD4 | PI | 12-3 | 27 | 24-1 | D1 | 1-2 | C1 |

Next, we wished to determine whether the clonally expanded T-cells had TCRs specific for the injected antigen. Hence, from each participant we selected clonally expanded TCRs to re-express in an *in vitro* cell based system(21, 22). We based the decision on which sequences to select on the size of the clonal expansions (with CD8^+^ T-cells having the largest expansions) as well as including smaller expansions of memory CD4^+^ (maximum expansion of 6 cells) and Treg cells (maximum expansion of 3 cells).

We re-expressed the TCRs in a mouse T-cell hybridoma, 5KC(21), expressing and challenged them with C19-A3 GNP, gold nanoparticles (GNP) and whole proinsulin (PI). Of 22 TCRs remade we found five that were GNP-reactive, and six that were proinsulin-reactive. A T-cell clone with two β-chains and a single α-chain was specific only with one of the β-chains (EEASI-B:Bl-4-proinsulin specific; EEASI-B:Bl-5 unknown specificity) (Figure 3A, B and Table 1). Recognition of proinsulin and GNP were HLA-restricted as illustrated by clones EEASI-A:Bl3 and EEASI-C:Bl6 (Sup Fig 2).

**Figure 3.**
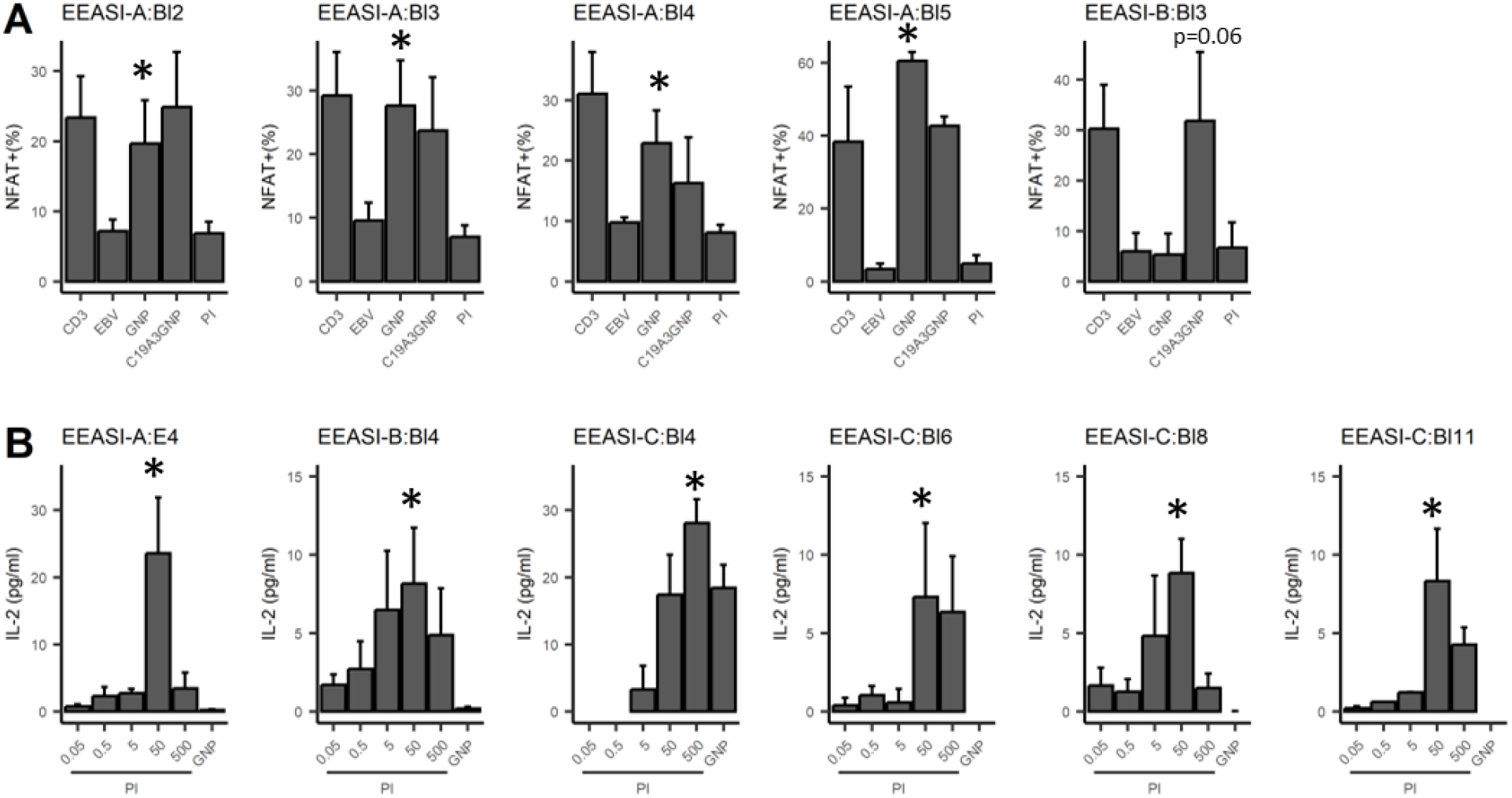
Antigen specificity of re-expressed TCRs. TCRs were re-expressed in 5KC cell lines. Each graph represents an individual 5KC cell line with a single TCR. A) GNP reactivity was measured by fluorescent reporter downstream of the NFAT promoter. CD3 - αCD3/CD28 positive control; EBV - patient-autologous EBV transformed cells used to present antigen-negative control; GNP: EBV transformed cells used to present GNP; C19-A3 GNP: EBV transformed cells used to present C19-A3 GNP; EBV transformed cells used to present PI - 50µg proinsulin. Data are pooled from three independent experiments. B) Proinsulin reactivity was assessed by mIL-2 production measured by ELISA, with background mIL-2 from 5KC+ EBV cells alone subtracted. GNP: 25µL GNP, PI: proinsulin concentration in µg/ml. Data are pooled from at least three independent experiments. Values indicate results of unpaired T-tests, *p<0.05

### Phenotype of clonally expanded T-cells differs depending on antigen-specificity

All the identified GNP-specific clones were CD8^+^, whilst PI specific clones were both CD8^+^ and CD4^+^. The majority of clones of unknown specificity were CD8^+^ with one exhibiting CD4^+^ Treg phenotype (Table 1, Figure 4). Additionally, many clonotypes had more than one phenotype. For example, some clones were found in both CD8^+^T_EM_ and CD8^+^T_RM/EM_ clusters, suggesting some plasticity of phenotype.

**Figure 4.**
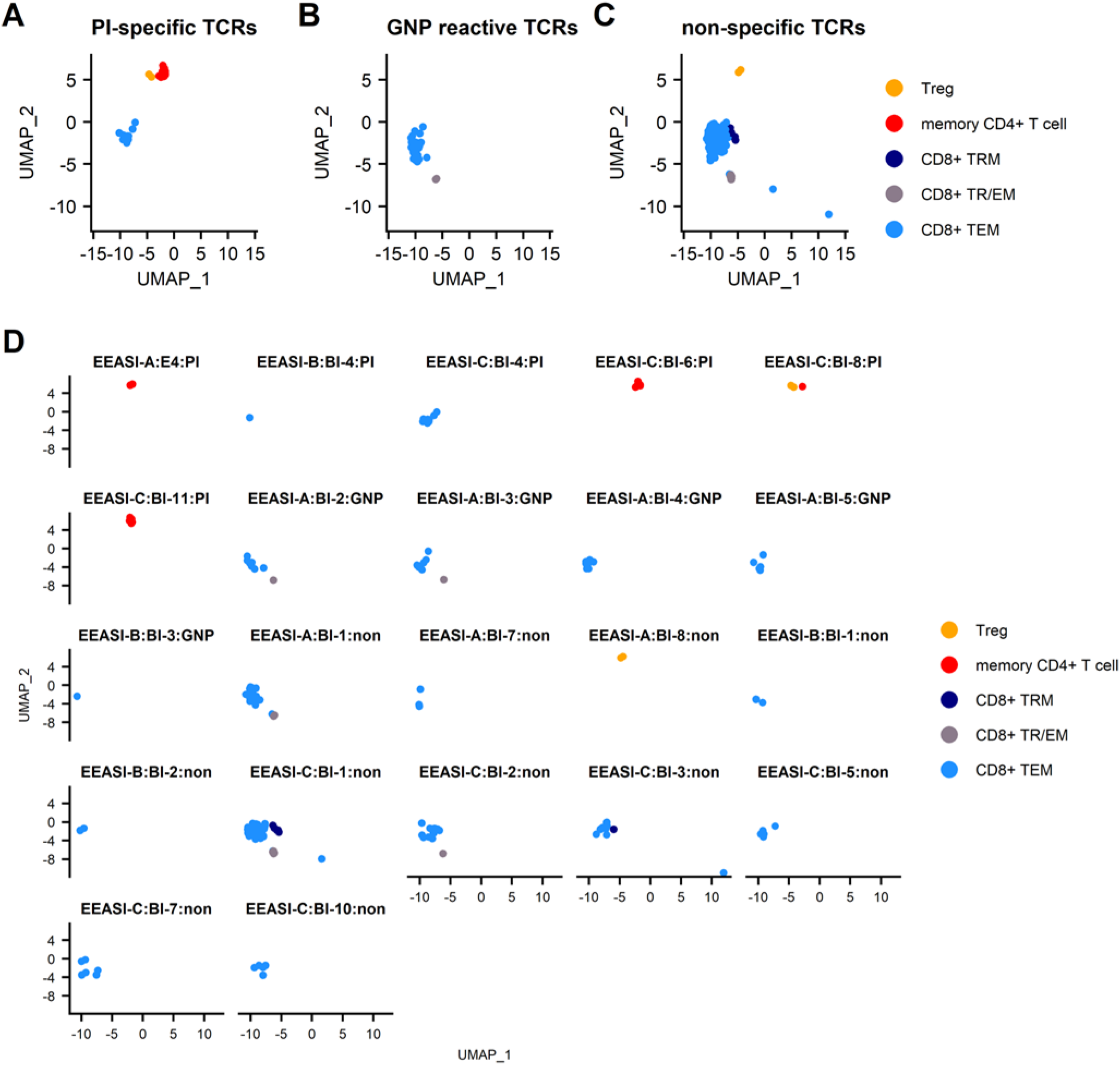
Specificities and T-cell phenotypes of clonally expanded cells in blister fluid. A) combined UMAP plot of PI-specific clones; B) combined UMAP plot of GNP-reactive clones; C) combined UMAP plot of clones of unknown specificity; D) UMAP plots of individual T-cell clones.

We next compared differentially expressed genes (DEG) between clones of unknown antigen specificity, GNP-reactive clones and proinsulin reactive clones (Supplementary Figure 3). As the majority of re-expressed TCRs from CD4^+^ T-cells were PI specific, it was not possible to compare differentially expressed genes between PI-specific and other specificity CD4^+^ T-cells. For CD8^+^ T-cells, Metascape pathway analysis between PI-specific and either gold-specific or non-specific T-cells revealed DEG in pathways associated with adaptive immune system activation and TCR signalling (Supplementary Figure 3). Examination of the top DEG revealed overexpression in PI-specific CD8^+^ T-cells of GNLY and KIR receptors. (Figure 5).

**Figure 5.**
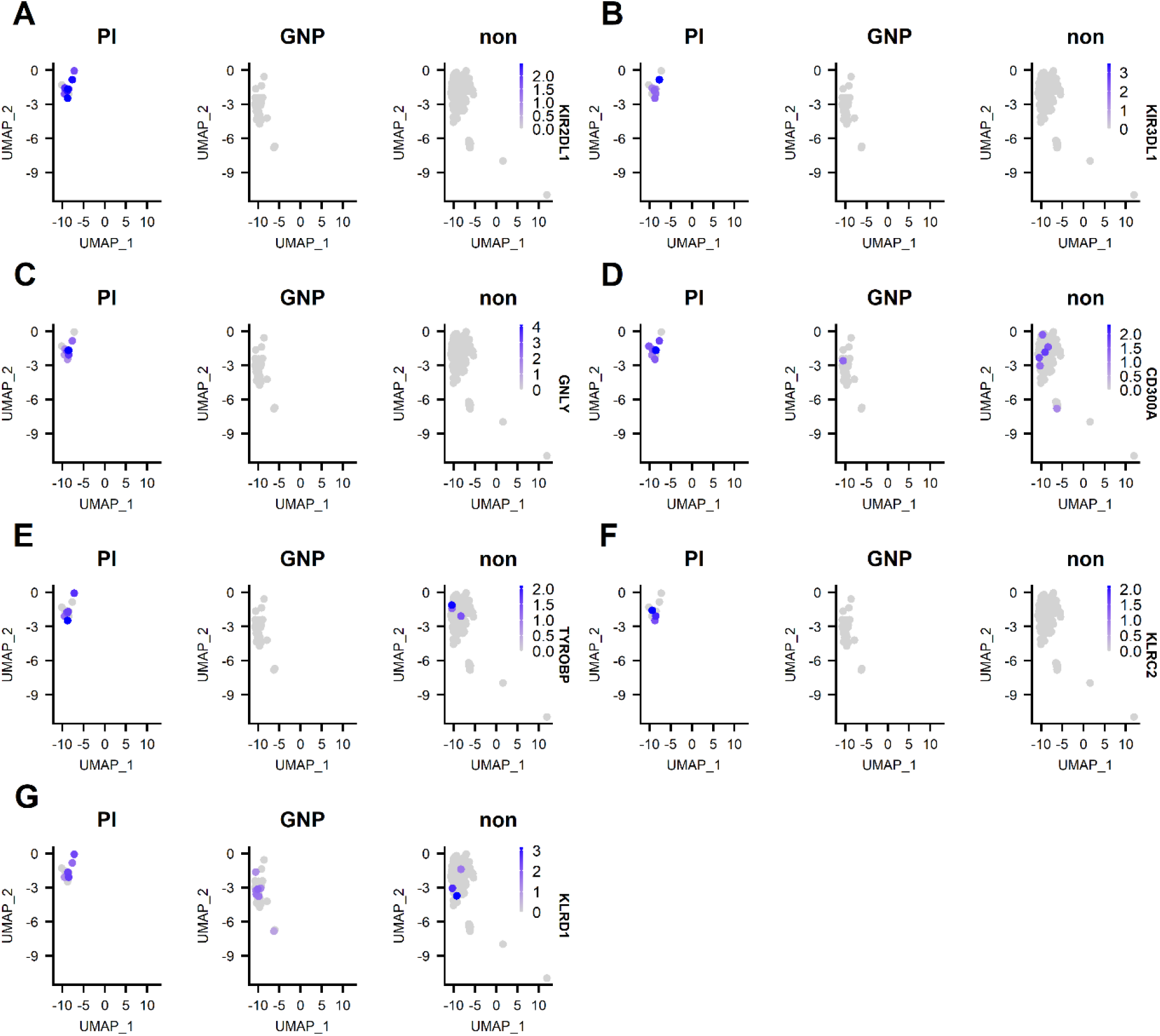
Top 5 DEG between PI-specific clones, gold-specific clones and clones of unknown specificity (designated non). A) expression of KIR2 DL1; B) expression of KIR3 DL1; C) expression of granulysin (GNLY); expression of KIR3 DL1; D) expression of CD300A; E) expression of TYROBP; F) KLRC2; G) KLRD1.

Despite the high level of specificity for the injected antigens, a limitation of the blister approach was a lack of healthy control individuals. Therefore, in order to determine whether the T-cells present in the blister had been recruited to the skin following injection of antigen, we next compared bulk RNA sequencing of punch biopsies from injection sites to those from healthy control individuals. We first searched canonical markers of leukocytes, T-cell subsets and chemokines and cytokines (Supplementary Figure 4). CD4^+^ and CD8^+^ transcripts were significantly upregulated in EEASI participants compared to control participants. The B cell marker CD19 was also upregulated, confirming the presence of B-cells in the confocal microscopy sections. Th1/CD8^-^ associated genes were particularly upregulated as were Th1-asscoiated chemokines and cytokines. Overall, DEG between T-cells from EEASI and control participants mapped to pathways primarily associated with immune cell activation (Supplementary Figure 4D). To directly correlate between the bulk RNAseq and scRNAseq we obtained a list of the top 10 genes that defined major cell types in the scRNAseq. We then searched for these genes within the punch biopsy RNAseq data and compared between control and EEASI samples. Nine out of 10 T-cell marker genes from the scRNAseq dataset were upregulated in EEASI compared to controls (Supplementary Figure 5A), as were all 10 markers for the CD8^+^T_EM_ subset (Supplementary Figure 5B).

## DISCUSSION

We have demonstrated that intradermal injection of GNP with proinsulin peptide antigenic cargo substantially enriched antigen-specific clonally expanded T-cells in the skin, compared to the frequencies found in the peripheral blood. Using scRNAseq of T-cells harvested from the fluid of blisters raised over the injection sites, we showed that the local T-cell response was directed to the gold core as well as the antigenic cargo. The antigen-specific cells were predominantly CD8^+^ T-cells with some CD4^+^ T-cells and had diverse phenotypes, which more resembled pancreatic islet T-cells than blood T-cells.

### Proinsulin-specific clonally expanded T-cells

We demonstrated reactivity of re-expressed TCRs to whole proinsulin rather than C19-A3 peptide, as responses to C19-A3 were generally weaker (data not shown). This is likely due to the efficacy of autologous EBV-transformed B cells to take up and process whole proteins vs peptide. However, we cannot exclude the possibility that the binding of the C19-A3 peptide to the gold directs its cleavage and presentation within the antigen presenting cells *in vivo*, favouring generation of immunogenic peptide fragments different from those produced *in vitro* from unconjugated C19-A3.

The finding of PI-specific CD8 clones is intriguing given that C19-A3 is known to be an HLA class II-restricted peptide(4, 23). Again, antigen processing to release smaller peptides suitable for HLA class I presentation is a possible explanation. There are limited reports in the literature of CD8^+^ T-cell epitopes within proinsulin C19-A3. Proinsulin C29-A4 is a reported CD8 epitope for HLA-A*02:01(24) (expressed by participant EEASI-C in our study(2)). HLA-B15 (expressed by both participants EEASI-B and EEASI-C) is reported to bind Proinsulin C29-A4, C30-A4 and C31-A4(25) (although tetramer assays for antigen-specific TCRs have not been reported), whilst proinsulin C23-32 is a HLA-A*03 epitope(26) (expressed by donor EEASI-B). A search of IEDB found no known epitopes within C19-A3 that would utilise donor EEASI-A HLA class I types, although many additional CD8 epitopes are known to reside within preproinsulin and to be directly involved in triggering the killing of β-cells(27, 28).

### Gold-specific clonally expanded T-cells and links to hypersensitivity

In addition to proinsulin specific clones, we identified GNP-specific CD8^+^ T-cell clones in the blister fluid and were able to demonstrate HLA-restricted reactivity. However, none of the re-expressed CD4^+^ TCRs were GNP-specific. As an element, gold is inert and unreactive, and requires ionisation to function as a hapten(29). This may have occurred due to the addition of linkers (glutathione and glucose) to the GNPs used in this study and the subsequent cleavage of the linkers either intracellularly or extracellularly.

Gold-reactive TCRs have previously been isolated from patients with rheumatoid arthritis who have received Gold sodium thiomalate(30). These TCRs were present in both CD4^+^ and CD8^+^ T-cells and some TCRVβ genes were overrepresented in gold-reactive TCRs. It has been suggested that gold reactivity is based on different mechanisms including covalent binding to large self-derived proteins (cell membrane proteins/MHC peptide complexes) when they are presented by APC, with or without processing. In other scenarios, metals can cross-link TCR to MHC independently of the nature of associated peptide. Gold may even bind and stimulate TCRs independently of MHC(30). However, others have found that gold-reactive TCRs require a compatible MHC(31).

Intriguingly in our study, all four tested participants exhibited a positive skin patch hypersensitivity test to gold thiosulphate as previously reported(2). Most cases of gold allergy involve delayed-type hypersensitivity(32), present in up to 15% of the population(33). It has been hypothesised that MHC-class I-independent CD8^+^ T-cell responses in response to gold-sensitised APCs is likely related to NKT-cell activation(34). However, in other metal allergies, conventional T-cells are implicated, in many cases with specific TCR sequences or V gene usages(35). We did not observe NKT-cells, and our clonally expanded CD8^+^ T-cells had a conventional phenotype. It is unclear how a positive gold skin patch test (of generally unclear clinical relevance (29, 36)) relates to the finding of gold-reactive clones in our study. Importantly, systemic hypersensitivity was not observed in any of the participants receiving C19-A3 GNP(2).

Finally, there were a significant number of clonal expansions of unknown antigen specificity. We speculate that these could be specific to other epitopes in the C19-A3 GNP compound or to unrelated antigens found in the skin. In addition, as responsiveness to antigens by TCR transduced cells is typically lower than that by primary T-cells, it is possible that specificity for PI or GNP was not detected in our assay system.

### Phenotype of the antigen-specific cells

We analysed the transcriptomes of T-cells found in the blister fluid of injection sites using bulk RNA seq (Supplementary Figure 4 and 5) and scRNAseq that captured all mRNA transcripts in each cell, allowing far deeper phenotyping compared with flow cytometry(16).

Traditionally, flow-cytometric approaches have focussed on analysing tetramer enriched autoantigen-specific T-cells from peripheral blood. For example, a recent study found that diabetes antigen-specific CD8+ T-cells from the peripheral blood of people with T1D displayed remarkably heterogenous phenotypes both within and between individuals, including naïve, memory and exhausted phenotypes(37). These findings echo earlier reports with more limited marker panels that found diabetes antigen-specific CD8^+^ T-cells in peripheral blood had diverse phenotypes spanning naïve, memory, effector and T_SCM_ phenotypes (although T_SCM_ were enriched in people with T1D compared to controls)(8).

In contrast, even in donors without diabetes, T-cells infiltrating the pancreas are dominated by CD8^+^ T-cells that display memory and effector memory phenotypes with markers of tissue residency(38). Others have reported that, in healthy donors, pancreatic T-cells are largely T_RM_, expressing CD127 (a marker of long-lived memory and antigen-independent replication), with no features of exhaustion and no CD57 expression, also expressing high levels of PD-1, GzmB, CD101, CD49a and CD103 compared to those in the peripheral lymph nodes (PLN) (39).

Pancreatic T-cells had highly expanded clonotypes, whereas those in the PLN had a greater diversity. Approximately 50% of clones in the pancreas were also found in the PLN(19, 39). These studies did not demonstrate antigen specificity of the T-cells. A comparison between islet-infiltrating cells in healthy donors and those with T1D found that CD8^+^T_RM_ cells were more abundant in people with diabetes. T_RM_ dominated the infiltrate (43% of CD8^+^ T-cells were CD69^+^CD103^+^) and there were relatively few effector CD8^+^ without features of tissue residency and memory(40). They found that the expression of genes associated with an acute cytotoxic response, such as *TNF, CCR7, LTA*, and *IL1A* or activated cytotoxic T-cells (*IL2R, CD40LG*, and *FASLG*), were not detected in any T1D subjects. However, genes encoding cytokines more related to inflammation and innate immune responses (eg, *IFNG, IL18, IL22,* and *IL15*), chemokines, and ligands *CXCL1, CCL11, CXCL9, CCL19, CCL7, CCL5*, and *CCL4* were up-regulated in insulitic islets of T1D subjects compared to non-insulitic islets from non-diabetic control donors(40).

We found that the cells in the blister had features along a spectrum of effector and memory phenotypes, with little evidence of naïve or exhausted phenotypes(8, 41, 42) (Supplementary Figure C and D). Only the CD8^+^ T_PM_ set showed evidence of naivety with expression of CCR7, CD62L and IL7R (Supplementary Figure C). Whilst only the CD8^+^ T_RM/EM_ population was actively proliferating (Ki67^+^), no subsets had substantial expression of CD57. Although many subsets expressed CD27, none had a CD27^+^CD95^+^CCR7^+^ phenotype characteristic of T_SCM_(8). Whilst we were not able to assess a functional response in our CD8^+^ T-cells, they were transcriptionally active for a number of cytotoxic transcripts, including GzmB, perforin and GNLY, suggesting that they were participating in an ongoing immune reaction and are more homologous to those found in the islets than normal skin resident CD8^+^ T-cells. Indeed, previous research into skin resident CD8^+^ T-cells has indicated that they are phenotypically distinct from those in the peripheral blood and lack immediate cytotoxic functions(43). In contrast, a cytotoxic phenotype was observed in our PI-specific CD8^+^ clones, as evidenced by DEG analysis, which revealed overexpression in GNLY and KIR receptors in comparison to gold-specific and clones of unknown specificity. KIRs interact with HLA molecules and KIRs with DL tails express immunoreceptor tyrosine-based inhibitory motifs. Significant differences in KIR gene DEG expression is an important finding, as polymorphism in those genes has been associated with T1D, with some genes being identified as protective and others as susceptibility factors for T1D(44–51). For example, children with T1D have fewer inhibitory KIRs with their corresponding ligands compared with healthy control children(50). As the expression of inhibitory KIRs is generally negatively associated with T1D, it is intriguing to see these overrepresented in the PI-specific CD8^+^ T-cells. However, it is known that the specificity of KIR expressed on CD8^+^ T-cells is random and often distinct compared with those expressed on NK cells within the same individual(44) and that KIR2DL1 expressing CD8^+^ T-cells have enhanced cytotoxic capabilities(45). Indeed, PI-specific clones consistently overexpressed KIR2DL1 in comparison to gold-specific and clones of unknown specificity. Whilst the number of clones included in this analysis was limited, it demonstrates an important proof of principle of how *in vivo* activation and enrichment can be combined with scRNAseq to phenotype diabetes autoantigen-specific T-cells. Although some blister clones showed regulatory properties, this was not the predominant phenotype.

It is open to speculation if the blister clones reactive to injected antigens derived from naive T-cells following antigen stimulation or arrived in the skin from a pre-existing memory pool following antigen stimulation. The appearance of the skin induration approximately 2 weeks after the first injection, but subsequently a much faster skin reaction after the 2^nd^ and 3^rd^ (1-3 days), potentially indicates that the cells have at least proliferated *in situ*. The presence of gold-reactive clones also supports this. Interestingly, we observed different phenotypes with the same clonotype indicating the transformation from effector to long-lasting tissue resident phenotype as a consequence of chronic antigen exposure(52, 53). Our data demonstrate that the phenotypes of the infiltrating T-cell population and more specifically, PI-reactive T-cells, found in blister fluid following C19-A3 GNP injection, more closely resembled the phenotypes of antigen-specific T-cells isolated from human pancreatic islets (Supplementary Figure 1 E,F) (54) than those isolated from blood of people with T1D.

A limitation of the study is the small number of participants that emphasises individual differences in immune response. In addition, post-injection timing of the samples had a wide range from 3-7 months for biopsies and 10-28 months for suction blisters. Therefore, comparisons are made between different stages of the immunological response to C19-A3 GNP, although late sampling time-points in all individuals make it more likely that processes were in the stage of tissue residency in all participants.

### Prolonged skin retention of clonally expanded cells and potential for application in clinical practice

Antigen-specific cells remained enriched and *in situ* for 10-28 months post treatment with C19-A3 GNP. It is likely that injection and chronic skin retention of this compound induced the conversion and proliferation of naive antigen-specific T-cells to a cytotoxic tissue-resident phenotype rather than recirculation of pre-existing memory cells to the skin. Long-term retention in the skin opens the possibility of a novel opportunity for sustained immune modulation using GNPs to deliver antigens. Further work is required to optimise compound characteristics to favour tolerogenic properties, essential for autoimmune diseases. For example, we have previously shown that minimally invasive intradermal delivery of diabetes autoantigens and pre-treatment of skin with steroids promotes tolerance for treatment of T1D(55–57). However, the system maybe already suitable in its current form for delivering traditional immune-stimulating vaccines. This is because this platform includes both intradermal delivery and extended duration of antigen presentation known to promote generation of T_RM,_ important for vaccine effectiveness(53).

In summary, by using novel scRNAseq technology we have shown that intradermal administration of peptide-gold nanoparticle resulted in recruitment and clonal expansion of antigen-specific T-cells. This opens a wide range of possibilities for application of this methodology in clinical practice.

## METHODS

### Study design

As described before(2), the Enhanced Epidermal Antigen Specific Immunotherapy Trial -1 (EEASI) study was a two-centre, open-label, uncontrolled, single-group first-in-human Phase 1A safety study of C19-A3 GNP peptide in individuals with T1D (https://clinicaltrials.gov/ct2/show/NCT02837094). The investigational medicinal product (IMP) was C19-A3 GNP (Midacore™), which comprises Midacore™ GNPs (Midatech Pharma Plc, Cardiff, UK)(1, 58) of a size of less than 5 nm, covalently coupled to an 18-amino acid human peptide, the sequence of which is identical to the residues from position 19 in the C-peptide of proinsulin through to position 3 on the A-chain of the same molecule (GSLQPLALEGSLQKRGIV). The peptide is synthesised with a linker to facilitate binding to the GNPs: 3-mercaptopropionyl-SLQPLALEGSLQKRGIV 2 acetate salt (disulfide bond). The chemical composition of the IMP contained a ratio of 4 C19-A3 peptides: 11 glucose C2: 29 glutathione ligands as determined by 1H-NMR (proton nuclear magnetic resonance). A typical batch contained: [C19-A3 peptide] = 1.33 mg/ml; [gold] = 5.5 mg/ml; [glucose linker] = 0.6 mg/ml; [glutathione linker] = 1.79 mg/ml. As the drug substance was diluted 1:7 to 1:10, depending on the content of C19-A3 peptide per particle, and as 50 μl of the diluted solution was administered to the study participants, this corresponded to C19-A3 peptide: 10 μg; gold: 39 μg; glucose linker: 4.3 μg; glutathione: 12.7 μg.

Participants diagnosed with T1D and confirmed to possess the *HLA-DRB1*∗*0401* genotype, were given three doses of C19-A3 GNP at 4-weekly intervals (weeks 0, 4, and 8) in the deltoid region of alternate arms (2 doses in one arm and 1 dose in the other arm) via CE-marked 600 μm length MicronJet600™ hollow microneedles (NanoPass Technologies Ltd. Israel) attached to a standard Luer-lock syringe. The single-dose given in 50 μl volume was equivalent to 10 μg of C19-A3 peptide.

The planned duration of the study was initially 20 weeks, but this was later extended to 52 weeks to continue safety follow-up of the participants due to a persistent local response to the intradermal injection (as described below). Skin changes at the injection site were followed up beyond 52 weeks in some individuals (12-28 months) and additional analyses were performed to determine the nature of these changes, which are the focus of this manuscript. Data on safety and metabolic effects following C19-A3 GNP administration has been published separately(2). Study participants are denoted by the prefix EEASI, followed by their study number in the results section.

### Blood samples

PBMCs were isolated on Ficoll–Paque Plus (1.077 6 0.001 g/ml, +20 °C; GE Healthcare Biosciences, Sweden) gradient centrifugation from the peripheral blood of participants. Samples were cryopreserved in CryoStor® (Sigma-Aldrich, Gillingham, UK) on the day of the collection and thawed on the day of the experiment.

### Suction blisters

Skin suction blisters were raised as described previously(55). Briefly, suction cups were applied to the deltoid region of participants’ arms, at the site of previous injection. Skin suction blisters were performed by gradually applying negative pressure (up to 60 kPa) from a suction pump machine VP25 (Eschmann, Lancing, UK) through a suction blister cup with a 15-mm hole in the base (UHBristol NHS Foundation Trust Medical Engineering Department, Bristol, UK) for 2–4 hr until a unilocular blister had formed within the cup. The cup was left in place for a further 30-60 minutes to encourage migration of lymphocytes into the blister fluid. The cup was then removed, and the blister fluid aspirated using a needle and syringe. Blister fluid was immediately suspended in 10ml heparinised RPMI (Gibco)+ 5% foetal calf serum Cells were counted then washed once and resuspended in an appropriate volume for scRNAseq. Samples were used fresh on the day of the collection.

### Punch biopsies

Punch skin biopsies of the local area were performed under aseptic conditions and under local anaesthetic (Lidocaine Hydrochloride Injection BP 2%, w/v), using a 6-mm sterile disposable biopsy punch. Following the biopsy, samples were divided with half immediately placed in 10% formalin and transported to the laboratory and the other half placed in RNA later^TM^ (Fisher Scientific, Loughborough, UK) for bulk RNA sequencing.

Control skin samples were obtained from female patients aged 19–82 years, following mastectomy or breast reduction after informed consent. Skin without obvious pathological findings, which was surplus to diagnostic histopathology requirements, was used in the experiments. Control punch biopsy samples were treated in the same manner as described above.

### Immunofluorescence confocal microscopy

4mm sections of formalin-fixed paraffin-embedded skin biopsies were examined via immunofluorescence protocols. In brief, sections were dewaxed with Histoclear^TM^ (Fisher Scientific, UK), rehydrated in an ethanol series (100%,90%,70%,50% and ddH_2_O) and underwent dual antigen retrieval protocols (namely, heated under pressure for 20mins in 10mM Citrate (pH6 buffer) and 20mins in 1mM EDTA (pH8)). Sections were then blocked with 5% normal goat-serum (Vector, UK), and serially stained with multiple combinations of antibodies specific for CD8, CD20 and Ki67 (Dako: 1:50 (#M7103); 1:700 (#M0755); and 1:400 (#M7240), respectively), and FoxP3 and CD4 (Abcam: 1:80 (#ab133616); CD4 (1:100; #ab20034, respectively) – each being incubated at 4°C overnight and employing tyramide signal amplification secondary reagents (AK-5002&AK5001; Vector, UK) to reveal the immune maker staining. Each antigen was examined in at least 2 complementary combinations to ensure comprehensive understanding of expression and for quality control (eg CD8 was examined in the following combinations: CD8/CD103/Ki67; CD8/CD103/FoxP3; CD8/CD103/CD11c; CD8/CD4/FoxP3 and Ki67 was examined CD20/CD4/Ki67 and CD8/CD103/Ki67. Imaging was carried out using an SP8 DMi8 Confocal Laser scanning microscope, where a minimum of 5 regions were imaged for each combination in every section to capture an accurate overview of the whole biopsy. Images were analysed and mean numbers of relevant immunostained cells were captured using Halo^TM^ Highplex module (Indica Labs, USA). Cell counts verified by manual counts in 25% of the images by two independent researchers in a blinded manner.

### RNA sequencing of punch biopsies

After cutting skin samples into small pieces, samples were homogenised and lysed by using Tissue Raptor II and QIAzol® Lysis protocol (QIAzol® Handbook 2021, QIAgen, Crawley, UK). The quality of RNA was routinely assessed by determining the A260:A280 ratio using NanoDrop2000 (Thermo Scientific, UK). RNA libraries were created using TruSeq stranded Total RNA with ribozero GOLD (illumina, UK) and sequenced on an illumina HiSeq 2500 platform with 2 x 75 bp reads.

### scRNAseq

Samples were processed using a 10xGenomics V1 human 5’GEX kit and libraries were constructed following standard 10xGenomics protocols. Samples were pooled and sequenced on Illumina NextSeq and MiSeq. Demuxed samples were processed individually using the 10xGenomics CellRanger 3.0.2 pipeline and GRCh38-1.2.0. Data were further processed using the Seurat V3 package in R(59) and DoubletFinder(60) was used to remove doublets. TCR sequences and clone numbers were added to the Seurat object metadata slot. TCR clone numbers refer to data generated in the CellRanger pipeline, even if clones of the same cell did not meet gene expression quality control thresholds. Raw data contained red blood cells, but these were removed automatically in the quality control (QC) process due to low levels of unique molecular identifiers (UMIs) and low unique genes identified. Cell clusters were determined by Seurat, and the CD4^+^ T-cell cluster was manually divided in to Tregs and memory CD4^+^ T-cells.

### Antigen specificity experiments

TCR specificity was determined as previously described(22, 28). Briefly TCR α-chain and β-chain genes, connected by the porcine teschovirus-1 2A peptide gene, were engineered into a murine stem cell virus–based retroviral vector. T-cell transductants were generated by retroviral transduction of the 5KC murine T-cell hybridoma cells lacking endogenous TCR expression and expressing human CD4 or CD8 according to the original T-cell phenotype. The 5KC cells also incorporate an NFAT-binding sequence followed by the fluorescent ZsGreen-1 gene, such that activation of the cells can be detected by flow cytometry(29). 5KC cells were spin-infected with replication-incompetent retroviruses that were produced from Phoenix cells transfected with the retroviral vectors and the pCL-Eco packaging vector. Cells transduced with retroviruses were then selected by sorting for CD3 expression. (CD3ε MicroBead Kit, mouse, Miltenyi Biotec).

5KC cells bearing TCRs were incubated with autologous EBV-transformed B cells with antigens or blocking antibodies as indicated at 1×10^6^ 5KC/ml and 1×10^6^ EBV-transformed B cells/ml in 200µL RPMI 1640 (Gibco)+penicillin/streptomycin (Gibco) +10% foetal calf serum (BioSera)+ 2-Mercaptoethanol (Thermo Fisher Scientific) in 96-well round bottomed plates. The positive control was 5µL αCD3/CD28 microbeads (Dynabead). The proinsulin (PI) used in these studies was a recombinant human PI and was a kind gift from Luciano Vilela, Biomm S.A., Brazil. The recombinant human PI has the same structure and amino acid sequence as that of the native human PI, but also incorporates a histidine affinity tag (sequence from N′ to C′: MAHHHHHHMGR), resulting in a total of 97 amino acids and a molecular weight of approximately 12,500 g/mol (personal communication Luciano Vilela)(57). C19-A3 GNP (Midacore™) has been previously described(2). Gold nanoparticles 5nm diameter were from Merck Life Sciences. Ultra-LEAF Purified anti-human HLA-DR Antibody [Clone: L243] and Ultra-LEAF Purified anti-human HLA-A,B,C Antibody [Clone: W6/32] were from Biolegend and used at a 1:800 dilution in cell culture. Flow cytometry was performed to evaluate the expression of NFAT-reporter fluorescent protein on a BD Canto II after 24h. Murine IL-2 ELISAs were performed on undiluted cell culture media following the manufacturer’s instructions (Biolegend) after 48h.

### Statistics

Plots and Statistics were carried out in GraphPad Prism version 9.5.0 for analysis of confocal microscopy images.

Unpaired T tests with FDR correction were used to compare gene expression of specific markers in punch biopsies between healthy controls and clinical trial participants. DEG pathway analysis was conducted using Metascape standard parameters(61) on all DEG with an FDR less than 0.05

DEGs that defined cell clusters in Seurat were determined using Seurat’s inbuilt statistical methods using the Wilcoxon rank sum test. DEG pathway analysis was conducted using Metascape standard parameters(61) on all DEG with an uncorrected p value less than 0.05.

### Ethics statement

This study was carried out with the approval of the UK Research Ethics Service, the Linköping Regional Research Ethical Review Board (RERB), the UK Medicines and Healthcare products Regulatory Agency (MHRA) for Clinical Trial Authorisation, and the Swedish Medical Products Agency (MPA). Written informed consent was obtained from all participants. The trial was conducted in compliance with the principles of the Declaration of Helsinki (1996) and the principles of Good Clinical Practice and in accordance with all applicable regulatory requirements including but not limited to the Research Governance Framework and the Medicines for Human Use (Clinical Trial) Regulations 2004, as amended in 2006. Further details are available at https://clinicaltrials.gov/.

### Data availability

scRNAseq and bulk sequencing data will be deposited in a publicly available repository at the point of publication.

## AUTHOR CONTRIBUTIONS

SJH - designing research studies, conducting experiments, acquiring data, analyzing data, writing the manuscript

TCT - designing research studies, conducting experiments, acquiring data, analyzing data EJSR - conducting experiments, acquiring data, analyzing data

NNV - conducting experiments, acquiring data

NW - conducting experiments, acquiring data, analyzing data LL - designing research studies, conducting experiments

RA - conducting experiments, acquiring data, analyzing data, writing the manuscript QZS - analyzing data

PL - conducting experiments, acquiring data, analyzing data, writing the manuscript RW - conducting experiments, acquiring data, analyzing data

MAMcA - designing research studies, conducting experiments, writing the manuscript MN - designing research studies, writing the manuscript

FSW - designing research studies, writing the manuscript

JHMY - designing research studies, acquiring data, analyzing data TT - designing research studies, analyzing data

JL - designing research studies, analyzing data, writing the manuscript CMD - designing research studies, analyzing data, writing the manuscript

DT - designing research studies, conducting experiments, acquiring data, analyzing data, writing the manuscript

## Data Availability

All data produced in the present study are available upon reasonable request to the authors

## ACKNOWLEDGMENTS

Funding: EE-ASI was a European research network (Collaborative Project) supported by the European Commission under the Health Cooperation Work Programme of the 7th Framework Programme, under the Grant Agreement 305305. SJH is funded by the Diabetes Research and Wellness Foundation Professor David Matthews Non-Clinical Research Fellowship. PL is funded by the Diabetes UK RD Lawrence Fellowship. EJSR is funded by a grant from the Leona M. and Harry B. Helmsley Charitable Trust. QZS was funded by a Society for Endocrinology summer studentship.

## SUPPLEMENTARY FIGURES

**Supplementary Figure 1.**
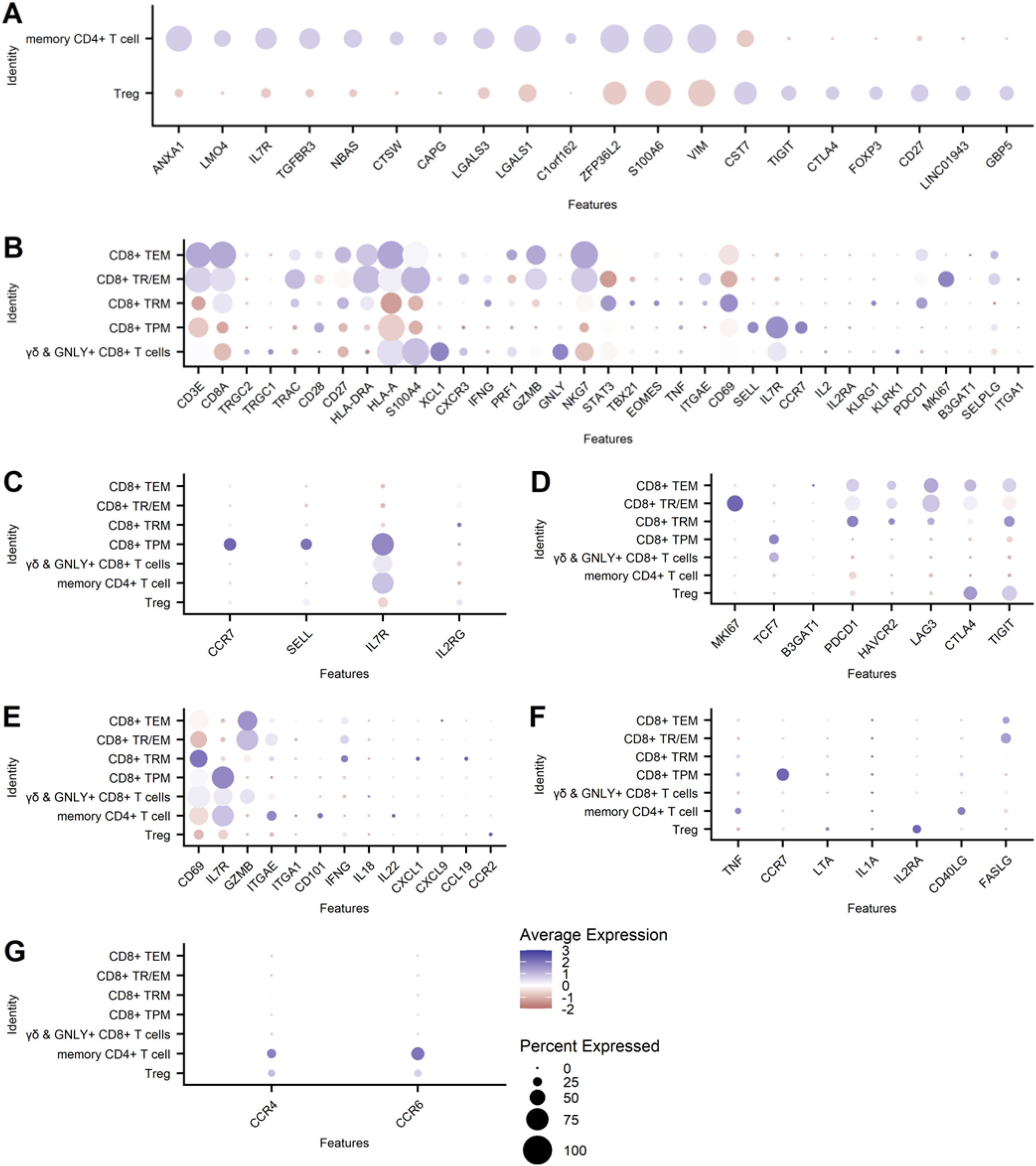
Phenotypes of cell subsets presented in dot plots: A)CD4_+_ cells were manually gated into two subsets and their identities were determined to be Tregs and memory CD4_+_ T-cells based on the top 20 DEG between the two subsets; B) Key markers of CD8_+_ T-cell subset identified based on canonical markers expression of memory, resident memory, peripheral memory and effector subsets and those found in the skin(43); C) naïve cell markers; D) exhaustion and proliferation markers in the T-cells; E) expression of genes known to be highly expressed in pancreatic resident T-cells; F) expression of genes known to have low expression in pancreatic resident T-cells; G) expression of skin homing chemokine receptors.

**Supplementary Figure 2:**
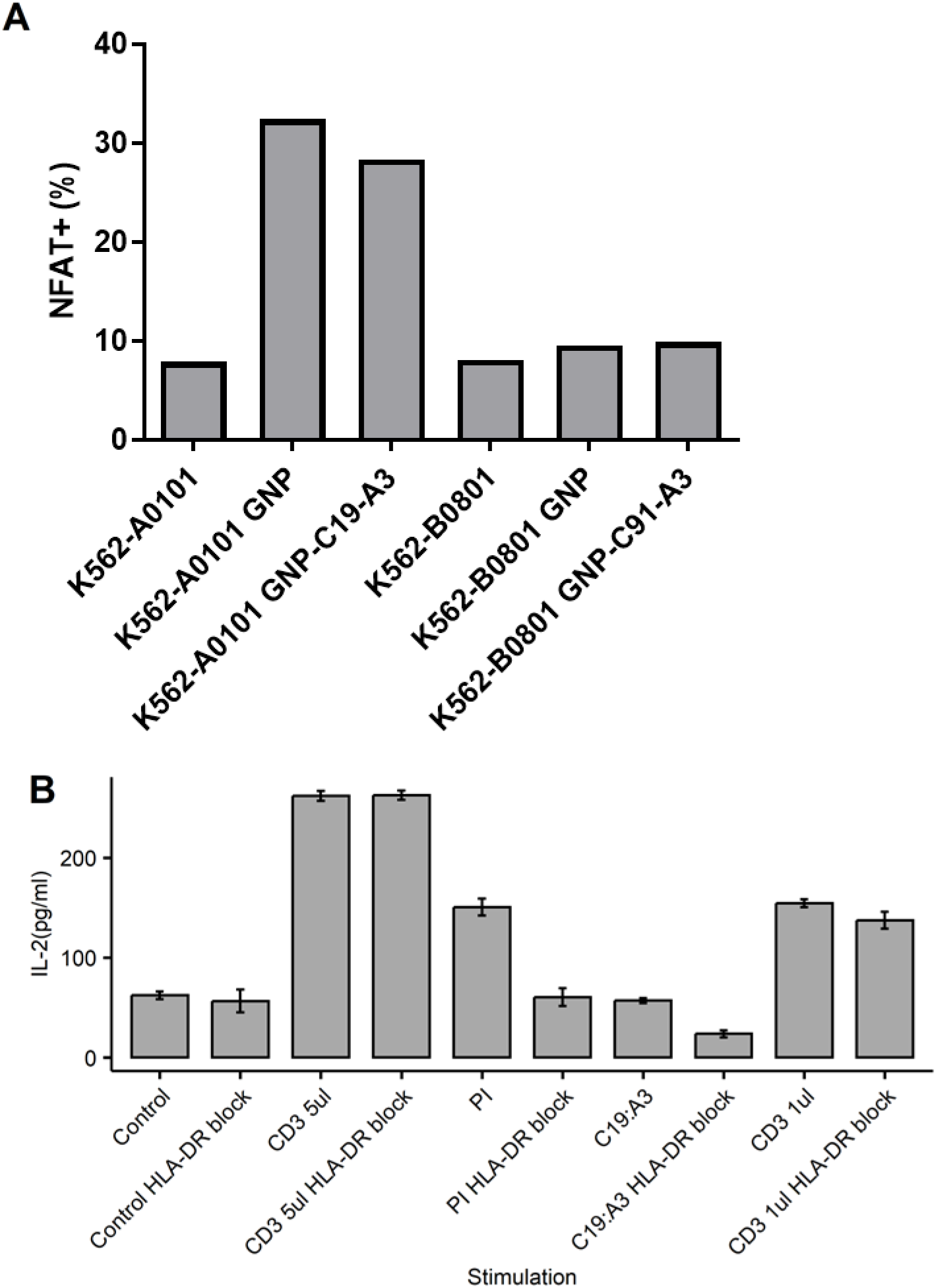
Example HLA restriction in GNP and PI-reactive TCRs. A) Clone EEASI-A:Bl3 is HLA A01:01 restricted. 5KC expressing the EEASI-A:Bl3 TCR were cultured with K562 cells that expressed either A01:01 or B08:01 (non-relevant HLA) either alone or with GNP or GNPC19-A3. Percentage fluorescence of the NFAT reporter was assessed after 24 hours by flow cytometry. B) Clone EEASI-C:Bl6 is HLA-DR restricted: Control - 5KC plus patient-autologous EBV transformed B cells; Control HLA-DR block – anti-HLA-DR blocking antibody present; CD3 5µL: 5 µL αCD3/CD28 microbeads present as positive control; PI:proinsulin 50µg/ml; C19:A3 – CD19-A3 peptide at 50µg/ml; CD3 1µL: 1µL αCD3/CD28 microbeads submaximal stimulation (not affected by HLA-DR block). HLA-DR block - Ultra-LEAF Purified anti-human HLA-DR Antibody at final concentration of 1.25µg/ml. Results are from single experiments representative of three independent experiments.

**Supplementary Figure 3.**
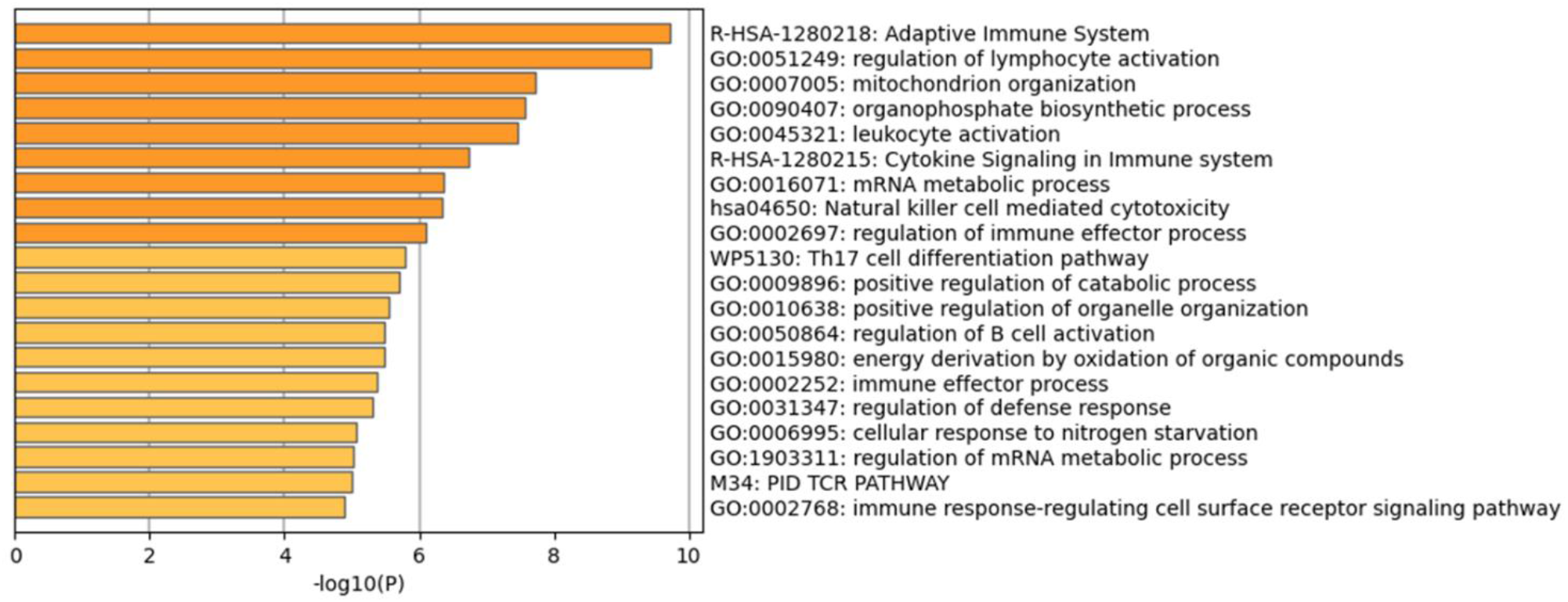
DEG between PI and GNP/non-specific T-cells in scRNAseq of blisters. DEG were identified using Seurat and all DEG with an uncorrected p value <0.05 were mapped to pathways in Metascape. The top 20 pathways are shown.

**Supplementary Figure 4.**
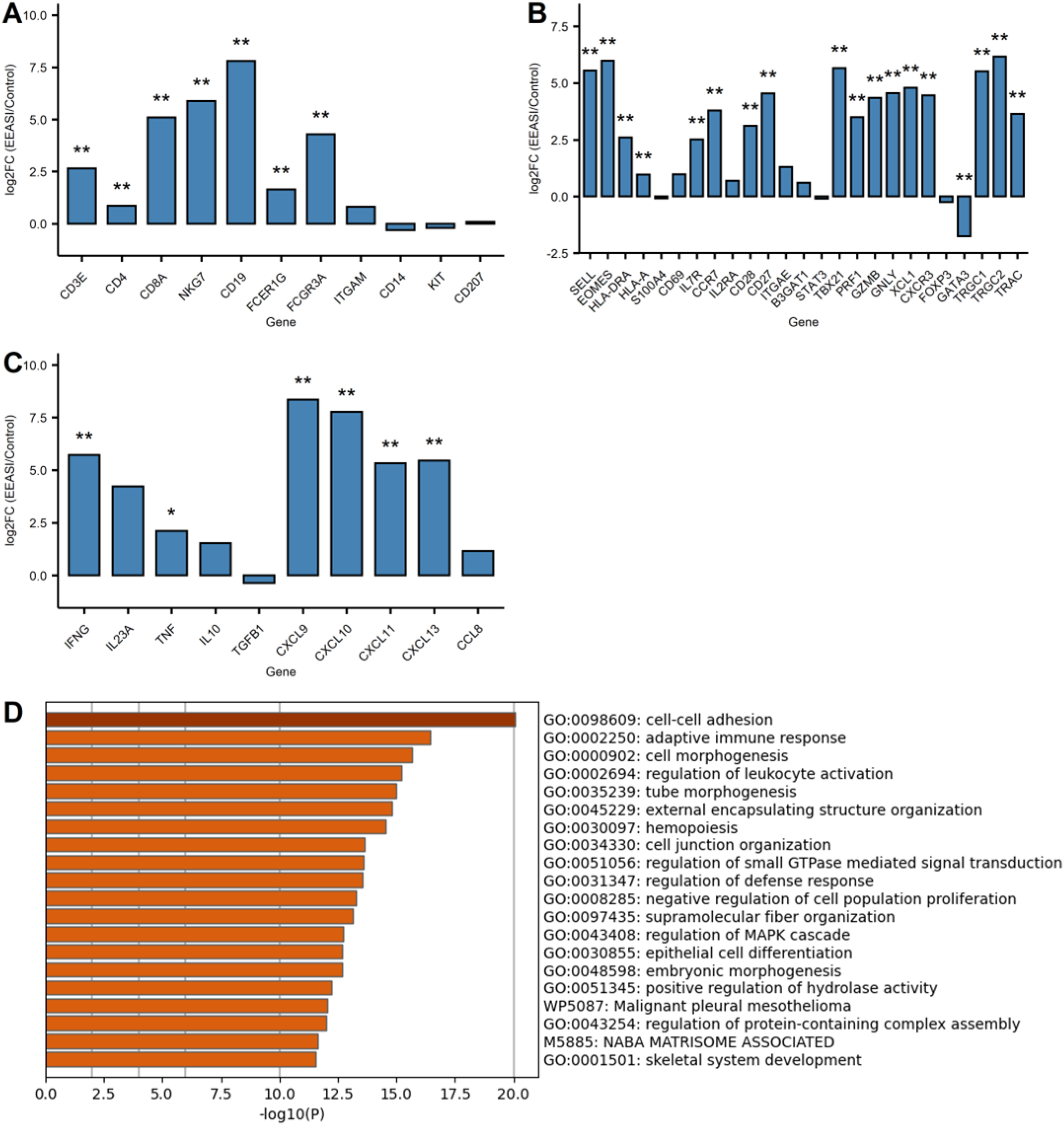
Immune cell and skin cell transcripts are differentially expressed between study participants and controls. Bulk RNAseq analysis was performed on punch biopsies from the injection site of three experimental study participants. Mastectomy skin samples, surplus to those required for diagnostic pathology, were used as controls. RNAseq analysis was performed to sequence all RNA transcripts and the log2 fold change (study participants/controls) in A) Leukocyte markers B) T-cell subset markers C) chemokines and cytokines. *p<0.05 **p<0.01 unpaired T test, FDR corrected d) All differentially expressed genes at p<0.05 FDR corrected, were mapped using Metascape analysis. The top 20 pathways are shown on the graph.

**Supplementary Figure 5.**
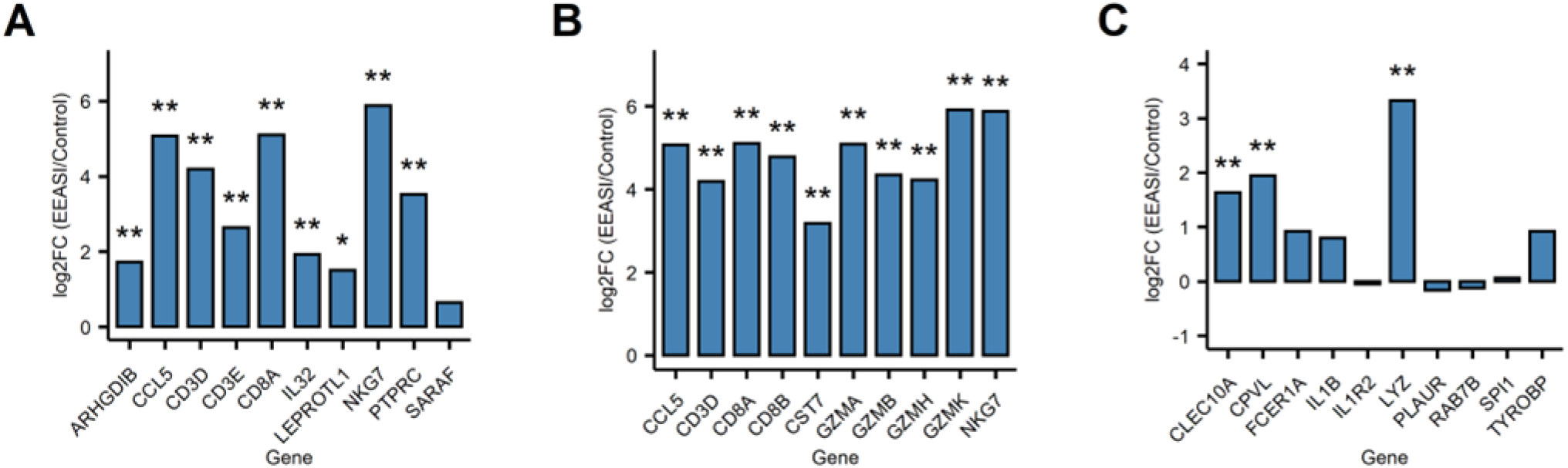
Immune cell and skin cell transcripts are differentially expressed between study participant and control based on marker genes from the scRNA seq. The top 10 DEG that defined A) all T-cells, B) CD8^+^ TEM and; C) DC were identified in the scRNAseq dataset. The relative expression of these genes between punch biopsy RNAseq between experimental study participants and control participants was then determined. *p<0.05, **p<0.01 unpaired T Test, FDR corrected.

